# Longer interval between maternal RSV vaccination and birth increases placental transfer efficiency

**DOI:** 10.1101/2024.07.14.24310390

**Authors:** Ms. Olyvia J. Jasset, Paola Andrea Lopez Zapana, Zeynep Bahadir, Lydia Shook, Ms. Maria Dennis, Ms. Emily Gilbert, Ms. Zhaojing Ariel Liu, Ms. Rachel V. Yinger, Ms. Caroline Bald, Ms. Caroline G. Bradford, Ms. Alexa H. Silfen, Sabra L. Klein, Andrew Pekosz, Sallie Permar, Liza Konnikova, Lael M. Yonker, Douglas Lauffenburger, Ashley Nelson, Michal A. Elovitz, Andrea G. Edlow

## Abstract

**Background:** Respiratory Syncytial Virus (RSV) is associated with significant neonatal and infant morbidity and mortality. Maternal bivalent RSVpreF RSV vaccination to protect neonates and infants was approved in September 2023 for administration between 32+0 and 36+6 weeks to protect neonates and infants. This approved timeframe is narrower than the 24-36 week window evaluated in the clinical trial, due to the possible association between preterm birth and vaccine administration. Currently, data are lacking on how maternal vaccine timing within the approved window affects the transfer of antibodies from mother to fetus, critical information that could influence clinical practice.

**Objectives:** We sought to examine how gestational age at vaccination and time elapsed from maternal RSV vaccination to delivery impacted transfer of maternal antibodies measured in the umbilical cord at delivery and in peripheral blood of 2-month infants. We also examined differences in maternal and cord RSV antibody levels achieved by vaccination versus natural RSV infection.

**Study Design:** A prospective cohort study was conducted at two academic medical centers between September 20, 2023 and March 21, 2024, enrolling 124 individuals who received the RSV vaccine during pregnancy. Infant capillary blood was collected at 2 months of age from 29 of the infants. Maternal and cord IgG levels achieved by RSV vaccination were compared to those associated with maternal natural RSV infection, using banked blood from 20 maternal:cord dyads collected prior to the availability of the maternal RSV vaccine. Levels of IgG against RSV strain A2 and B fusion (F) and attachment (G) proteins and against pertussis toxin (as a comparator antigen from a vaccine routinely administered earlier in pregnancy) were measured using a Binding Antibody Multiplex Assay. Differences in titers between vaccination and natural infection were examined using Wilcoxon rank sum test. Differences in cord:maternal transfer ratios and 2-month infant antibody levels by timing of maternal vaccination were evaluated by Kruskal-Wallis testing.

**Results:** Maternal RSV vaccination resulted in significantly higher maternal and cord anti-F RSV antibody levels than natural infection (5.72 vs 4.82 log_10_MFI, p < 0.0001 maternal; 5.81 vs 5.03 log_10_MFI, p < 0.0001 cord). Maternal vaccination 2-3 weeks and 3-4 weeks prior to delivery was associated with significantly lower cord:maternal transfer ratios than were observed when vaccination occurred > 5 weeks prior to delivery (p = 0.03 for 2-3 weeks, p = 0.007 for 3-4 weeks), and significantly lower transfer ratios than observed for pertussis vaccination administered prior to 30 weeks’ gestation (p = 0.008 for 2-3 weeks, p = 0.03 for 3-4 weeks, similar at > 4 weeks).

**Conclusion(s):** Vaccine administration earlier in the approved 32-36 week window (at least 5 weeks prior to delivery) results in the highest transplacental transfer of maternal antibodies to the neonate. These results should inform the counseling of pregnant individuals on optimal vaccination timing.

## Introduction

Respiratory Syncytial Virus (RSV) is a leading cause of infant respiratory disease and hospitalization globally, and the primary cause of infant hospitalization in the United States, affecting 2-3% of infants under 6 months of age.^1–4^ The highest morbidity occurs in preterm infants, who account for 25% of pediatric RSV hospitalizations.^5^ The MATISSE trial, a large randomized controlled trial of RSV vaccination, demonstrated that vaccination in pregnancy with the bivalent RSVpreF recombinant protein (Abrysvo, Pfizer) significantly reduced newborn and infant morbidity and mortality from RSV.^6^ In September 2023, the CDC recommended administration of this RSV vaccine to pregnant individuals.^4,7^ While the vaccine was administered between 24+0-36+6 weeks in the MATISSE trial,^6^ the approved gestational age for vaccination was limited to 32+0-36+6 weeks, due to concerns about vaccine-associated preterm birth.^7–9^ Data are lacking on the impact of timing of maternal vaccination within this window on transplacental transfer of maternal antibody, a correlate of infant protection.

The monoclonal antibody nirsevimab has been approved for administration to infants born within 14 days of maternal vaccination.^4,10,11^ However, both the American College of Obstetricians and Gynecologists and the CDC’s Advisory Committee on Immunization Practices (ACIP) acknowledge that this is an understudied area, with “<u>at least</u> 14 days…likely needed after maternal vaccination for development and transplacental transfer of maternal antibodies to protect the infant.”^4,10,12^ Detailed quantification of transplacental transfer efficiency of RSV-specific antibodies week-by-week after maternal vaccination could provide key data to inform the recommended window of administration of monoclonal antibody to neonates and infants. Moreover, this information can guide the future recommendations regarding the timing of maternal vaccination, particularly given recent real-world safety data suggesting no clear association between Abrysvo vaccine administration and preterm birth.^13^

Our prior work in the context of mRNA COVID-19 vaccines demonstrated that timing of maternal vaccination was associated with altered maternal responses and antibody transfer to the fetus,^14–18^ with administration at progressively later gestational ages associated with significantly reduced 6-month infant antibody levels.^18^ There is thus a concern that the narrowed gestational window for which Abrysvo was approved will similarly impact neonatal and infant immunity.

In the current hybrid model of prenatal care (a mix of remote and in-person visits) that arose during the COVID-19 pandemic, many pregnant individuals are seen in the office once at the beginning of the 32-36 week approved RSV vaccine window, and then not again until nearly 36 weeks. Currently, vaccination at all weeks across the 32+0 and 36+6 period is treated as equivalently protective for the neonate and infant. Here, we leveraged a diverse cohort of pregnant individuals and their infants to address this clinically-important question: How does gestational age at vaccine administration within the approved window, or time elapsed from vaccination to delivery, alter transplacental transfer of maternal anti-RSV antibody and durability in infant blood? We found that maternal vaccination > 5 weeks prior to delivery yielded the most efficient placental transfer of antibody to the infant.

## Materials and Methods

### Study population

Pregnant and postpartum individuals at two academic medical centers were approached for enrollment of themselves and their infants between September 20, 2023, and March 21, 2024. Eligible participants were: (1) pregnant or 2-months postpartum (2) at least 18 years old, (3) able to provide an informed consent, and (4) recipients of the Abrysvo RSV vaccine in pregnancy. Potential participants were identified via obstetric schedules, referred by practitioners at the participating hospitals, or self-referred. Banked samples from 20 maternal-cord dyads collected prior to the availability of the Abrysvo RSV vaccine (enrolled between March 15, 2021 – November 11, 2021) were used as comparator group to evaluate differences between the top quartile of RSV antibody responses elicited by natural infection^19^ versus those elicited by vaccination. All pregnant participants signed informed consent for sample collection (MGB IRB #s 2023P001874, 2020P003538, and MSSM IRB 23-01383), and birthing parents signed consent for their infants (MGB IRB #2020P000955).

124 vaccinated pregnant individuals were prospectively recruited for the study, with maternal and/or umbilical cord blood samples from N=122 pregnant individuals and N=29 of their 2-month infants included in the final analyses (**Figure 1** and **Table 1**), along with plasma from N=20 maternal:cord dyads recruited into our pregnancy biorepository prior to the availability of the RSV vaccine in pregnancy. All RSV vaccinated participants were included in the analysis, regardless of the level of RSF F antibodies present.

**Figure 1:**
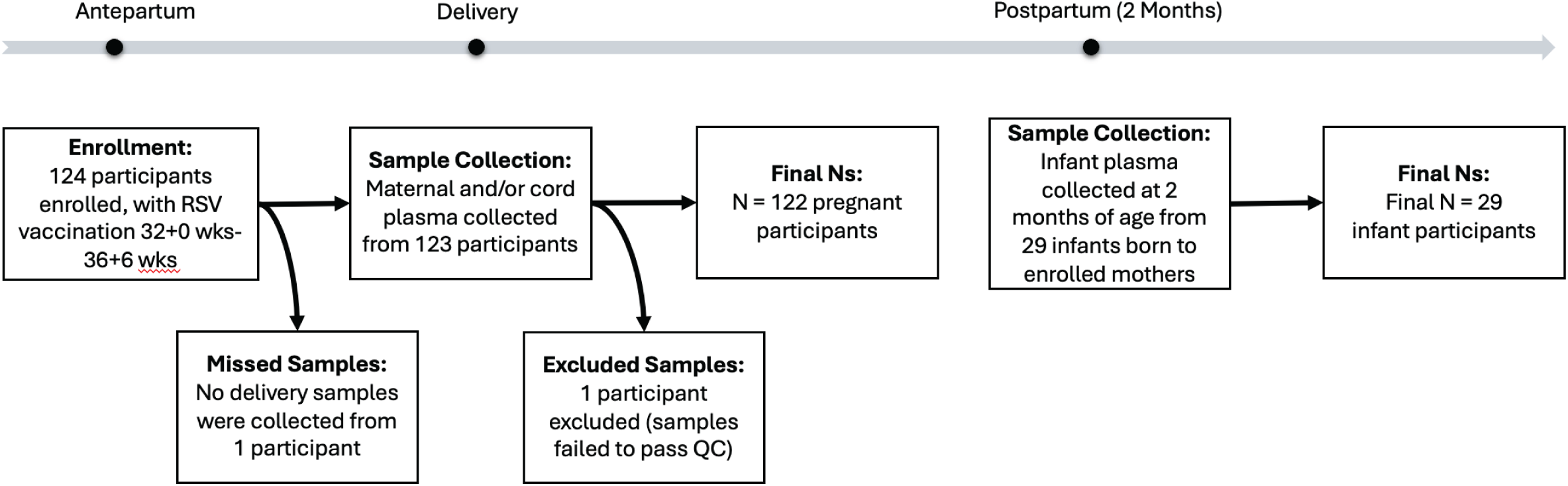
Flow diagram of participant enrollment, sample collection, and exclusion throughout the study.

**Table 1.** Participant demographics and clinical characteristics. Continuous variables are recorded as mean (SD); categorical variables are recorded as N (%).

| **Weeks between RSV Vaccination and Delivery** |  |  |  |  |  |  |
| --- | --- | --- | --- | --- | --- | --- |
| **Characteristic** | **All**<br>**N=142** | **RSV**<br>**Unvaccinated**<br>**N=20** | **< 2**<br>**weeks**<br>**N=4** | **2-4**<br>**weeks**<br>**N=21** | **4-6**<br>**weeks**<br>**N=49** | **> 6**<br>**weeks**<br>**N=48** |
| Maternal age (y),<br>mean (SD) | 34.7<br>(5.0) | 34.3 (4.4) | 39.0<br>(4.1) | 34.2<br>(5.9) | 35.6<br>(4.8) | 33.8<br>(4.9) |
| Race, N (%) |  |  |  |  |  |  |
| White | 99 (70) | 14 (70) | 1 (25) | 16 (76) | 35 (71) | 33 (69) |
| Black | 10 (7) | 0 (0) | 1 (25) | 1 (5) | 5 (10) | 3 (6) |
| Asian | 19 (13) | 5 (25) | 1 (25) | 2 (10) | 4 (8) | 7 (15) |
| Multiracial | 7 (5) | 0 (0) | 0 (0) | 1 (5) | 3 (6) | 3 (6) |
| Unknown | 7 (5) | 1 (5) | 1 (25) | 1 (5) | 2 (4) | 2 (4) |
| Ethnicity, N (%) |  |  |  |  |  |  |
| Hispanic or Latino | 16 (11) | 0 (0) | 0 (0) | 2 (10) | 7 (14) | 7 (15) |
| Not Hispanic or<br>Latino | 122 (86) | 19 (95) | 4 (100) | 17 (81) | 41 (84) | 41 (85) |
| Unknown | 4 (3) | 1 (5) | 0 (0) | 2 (10) | 1 (2) | 0 (0) |
| Public insurance | 25 (18) | 2 (10) | 0 (0) | 5 (24) | 7 (14) | 11 (23) |
| Gestational age at<br>RSV vaccination,<br>mean (SD) | 33.7<br>(1.4) | N/A | 34.9<br>(1.6) | 35.4<br>(1.2) | 33.8<br>(1.3) | 32.6<br>(0.6) |
| Nulliparous | 64 (45) | 10 (50) | 2 (50) | 12 (57) | 16 (33) | 24 (50) |
| Gestational age at<br>Delivery, mean (SD) | 39.1<br>(1.3) | 39.2 (1.1) | 35.9<br>(1.5) | 38.5<br>(1.3) | 38.8<br>(1.2) | 39.7<br>(0.8) |
| Preterm birth<br>(<37 weeks) | 8 (6) | 1 (5) | 3 (75) | 2 (10) | 2 (4) | 0 (0) |
| Infant female sex | 64 (45) | 8 (40) | 3 (75) | 10 (48) | 18 (37) | 25 (52) |
| Infant birthweight (grams), mean (SD) | 3352.5 (495.3) | 3416.3 (405.7) | 2555.0 (567.5) | 3202.4 (610.6) | 3406.3 (462.2) | 3403.0 (447.9) |
| Received Tdap | 140 (99) | 20 (100) | 4 (100) | 20 (95) | 48 (98) | 48 (100) |
| Gestational at Tdap, mean (SD) | 28.7 (1.7) | 28.0 (2.2) | 30.6 (3.1) | 28.8 (1.4) | 29.0 (1.8) | 28.6 (1.3) |
| **Included Infants** |  |  |  |  |  |  |
| Characteristic | N=29 | N=0 | N=1 | N=5 | N=14 | N=9 |
| Race, N (%) |  |  |  |  |  |  |
| White | 23 (79) | 0 (0) | 1 (100) | 5 (100) | 10 (71) | 7 (78) |
| Black | 1 (3) | 0 (0) | 0 (0) | 0 (0) | 1 (7) | 0 (0) |
| Asian | 3 (10) | 0 (0) | 0 (0) | 0 (0) | 1 (7) | 2 (22) |
| Unknown | 2 (7) | 0 (0) | 0 (0) | 0 (0) | 2 (14) | 0 (0) |
| Ethnicity |  |  |  |  |  |  |
| Hispanic or Latino | 2 (7) | 0 (0) | 0 (0) | 1 (20) | 1 (7) | 0 (0) |
| Not Hispanic or Latino | 25 (86) | 0 (0) | 1 (100) | 4 (80) | 11 (79) | 9 (100) |
| Unknown | 2 (7) | 0 (0) | 0 (0) | 0 (0) | 2 (14) | 0 (0) |
| Female sex | 16 (55) | 0 (0) | 1 (100) | 3 (60) | 8 (57) | 4 (44) |
| Birthweight (g), mean (SD) | 3376.4 (505.4) | 0.0 (0.0) | 2625.0 (0.0) | 3548.0 (753.9) | 3353.9 (467.0) | 3399.4 (405.9) |
| Received Nirsevimab | 1 (3) | 0 (0) | 1 (100) | 0 (0) | 0 (0) | 0 (0) |

### Sample collection, processing, and antibody quantification

Maternal and umbilical vein blood samples were collected by venipuncture into EDTA tubes (BD Vacutainer K3 EDTA, Becton Dickinson) at delivery admission and birth; respectively. Samples were collected between October 25, 2023 and April 4, 2024. Blood was centrifuged at 1000 g for 10 minutes at room temperature. Plasma was aliquoted into cryogenic vials and stored at −80°C until antibody quantification. Infant capillary plasma was collected via microneedle device (TAP II blood collection device, YourBio Health, Medford, MA) from January 19, 2024 – April 9, 2024.

IgG antibodies against RSV strain A2 and B fusion (F) and attachment (G) proteins and against pertussis toxin (a comparator antigen from a vaccine routinely administered earlier in pregnancy) were quantified using a Binding Antibody Multiplex Assay (BAMA) as previously described,^20,21^ and detailed in Supplementary Methods. All samples were run in duplicate, and %CV was evaluated with acceptance below 30%.

### Statistical Analysis

Statistical analyses were performed in R (v4.3.2). Luminex data was log_10_ transformed prior to analysis. For unpaired analysis comparing maternal, cord, and infant samples, or samples stratified based on the timing of the maternal RSV relative to delivery, a Kruskal Wallis test was performed, followed by Dunn’s post hoc test. P-values were further corrected for multiple comparisons using the Benjamini-Hochberg procedure. Unpaired analyses of two groups were performed using Wilcoxon rank-sum test. Paired analyses between the maternal and cord dyads were performed using Wilcoxon signed-rank test. Transfer ratios were calculated by dividing the cord plasma MFI value by the respective maternal plasma MFI value. The relative contributions of time from vaccination to delivery and gestational age at vaccination to transplacental transfer of maternal IgG were examined in a linear mixed effects regression model.

## Results

Vaccinated participants had significantly higher median anti-F RSV antibody levels than the upper quartile of unvaccinated participants in both maternal blood (5.72 vs 4.82 log_10_MFI, p < 0.0001) and cord blood (5.81 vs 5.03 log_10_MFI, p < 0.0001, **Figure 2A**). However, anti-G antibody levels were significantly higher in unvaccinated than vaccinated participants in both the maternal and cord blood (3.8 vs 4.2 log_10_MFI, p < 0.001 and 3.9 vs 4.3 log_10_MFI, p < 0.0001; respectively, **Figure 2B**), an expected result since the unvaccinated group represents the upper quartile of individuals with RSV antibodies. Similar results were identified for RSV B (**Figure 2**, bottom row). These results suggest that vaccination with the bivalent preF vaccine does not boost anti-G antibody levels in either maternal or cord blood, and confirm that the magnitude of F antibody levels are significantly greater after vaccination than those present in the top quartile of unvaccinated individuals.

**Figure 2:**
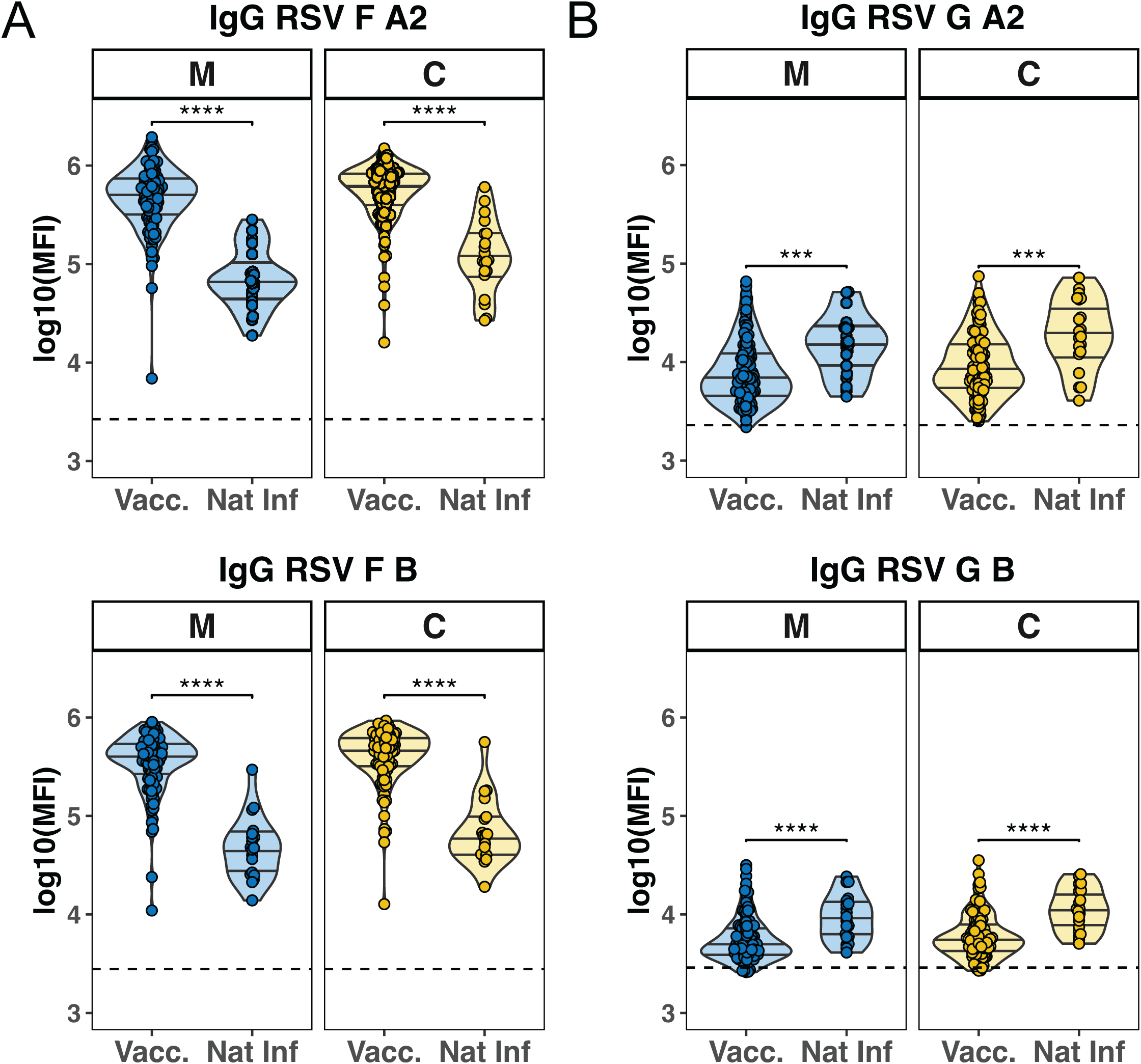
IgG antibody levels against RSV A2 and B Fusion and G proteins in vaccinated versus naturally-infected maternal:cord dyads. The violin plots show the log_10_ transformed levels of IgG antibodies targeted against (**A**) RSV A2 Fusion (**top**) and RSV B Fusion (**bottom**) in addition to (**B**) RSV A2 G antigen (**top**) and RSV B G Antigen (**bottom**) in N=117 vaccinated (Vacc.) maternal (M) and cord (C) samples, and N=20 naturally infected/unvaccinated (Nat Inf.) maternal and cord samples. The dashed line indicates the threshold of detectability, defined as the median + 3 standard deviations above PBS background levels. Differences between antibody levels in the vaccinated and naturally-infected/unvaccinated maternal and cord samples were assessed by Wilcoxon rank-sum test (\*\*\**p*<0.001, \*\*\*\**p*<0.0001).

Anti-F and anti-G RSV A/B antibody levels were quantified in triads of maternal, cord, and 2-month infant plasma (Figure 3). As expected in efficient transplacental transfer (cord:maternal ratios ≥1)^22,23^, levels of anti-F A2 RSV antibody were significantly higher in cord blood than in maternal blood (5.81 vs 5.72 log_10_MFI, p < 0.05). Anti-F A2 antibodies waned by 2 months of age, with significantly lower levels of anti-F A2 antibodies in 2-month infant plasma compared to both cord (5.35 vs 5.81 log_10_MFI, p < 0.0001) and maternal plasma (5.35 vs 5.72 log_10_MFI, p < 0.0001, **Figure 3A/Supplementary Figure 1A**). Anti-G RSV antibodies (a marker for RSV infection) also waned from birth to 2 months of age (3.92 log_10_MFI in cord blood vs 3.52 2-month infant, p < 0.0001, **Figure 3B/Supplementary Figure 1A**). Similar patterns were noted for RSV B strain (**Figure 3 bottom row/Supplementary Figure C**). An analysis of antibody persistence in 2-month-old infants using the area under the curve (AUC) of antibody levels from birth (umbilical cord) to 2 months of age demonstrated that anti-F RSV antibodies (boosted by vaccination) had significantly greater persistence than did anti-G RSV antibodies (presumably from prior infection, AUC value 0.7 vs 0.3, p < 0.0001) - **Supplementary** Figure 1B**/D**.

**Figure 3:**
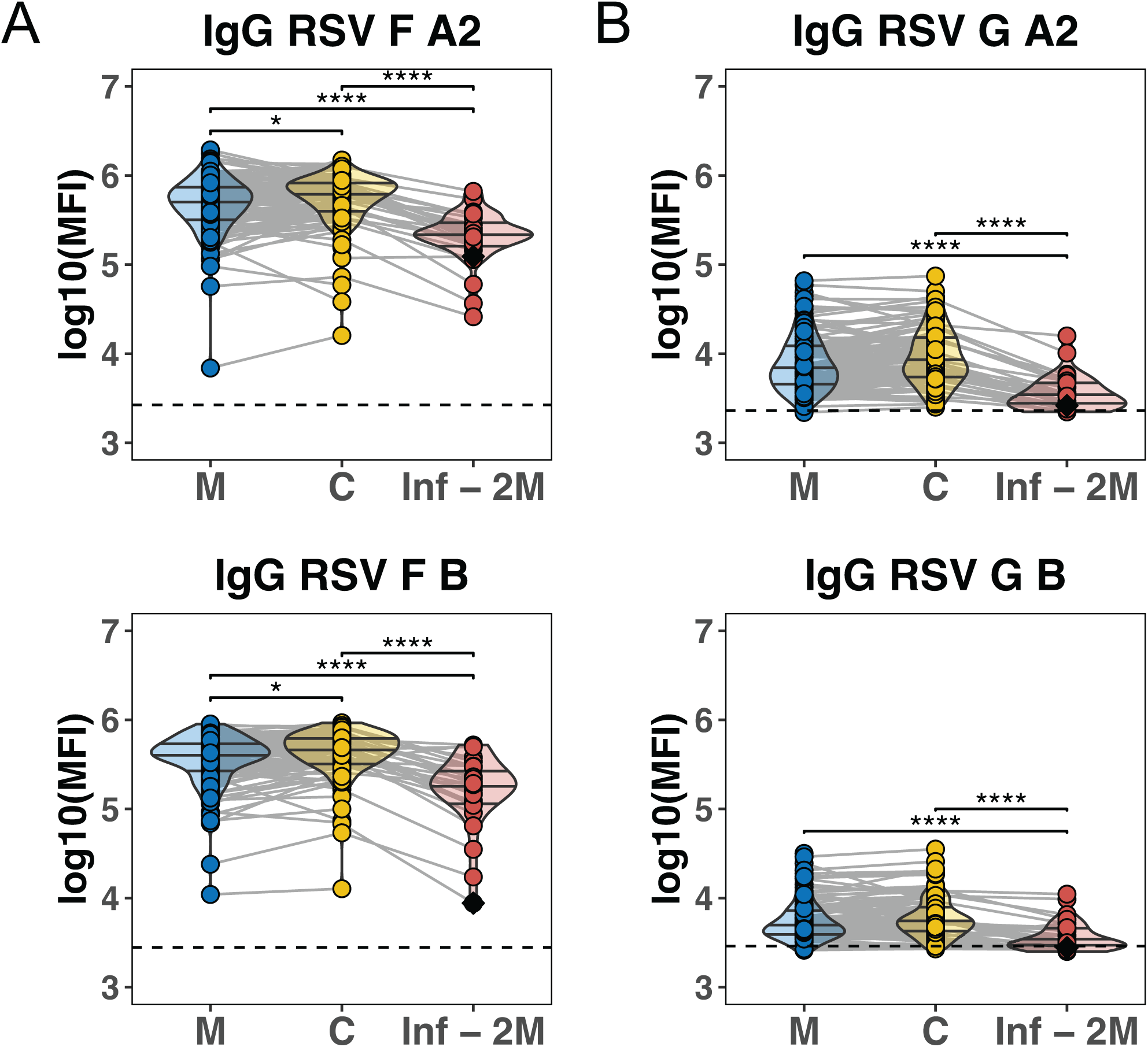
IgG antibodies against RSVA2 and B Fusion and G proteins in mothers at delivery, umbilical cord blood, and 2-month infants. The violin plots show the log_10_ transformed levels of IgG antibodies against **(A)** RSV F A2 **(top)** and RSV F B **(bottom)** and **(B)** RSV G A2 antigen **(top)** and RSV G B antigen **(bottom)** in N=117 vaccinated maternal (M) cord (C) dyads, and N=29 2-month infants (Inf -2m). The black diamond represents a sample from an infant who received Nirsevimab at 2 weeks old. The dashed line indicates the threshold of detectability, defined as the median + 3 standard deviations above PBS background levels. Differences between groups were assessed by Kruskal-Wallis test, followed by Dunn’s post hoc test when relevant. P values were further corrected using the Benjamini-Hochberg procedure (\**p*<0.05, \*\*\*\**p*<0.0001).

The transplacental transfer of maternal anti-RSV F IgG by weeks elapsed from vaccination to delivery was then evaluated using cord:maternal transfer efficiency ratios (Figure 4A). Vaccination 2-3 weeks prior to delivery was associated with significantly lower cord:maternal transfer ratios than were observed when vaccination occurred 5-6 weeks prior to delivery (0.76 vs 1.35, p = 0.03), or >6 weeks prior to delivery (0.76 vs 1.43, p = 0.008). Similarly, vaccination 3-4 weeks before delivery also resulted in lower cord:maternal transfer ratios than vaccination >6 weeks prior to delivery (0.92 vs.1.43, p = 0.03) indicating that vaccination earlier in the approved 32-36 week window, at least 5 weeks prior to delivery, results in significantly more efficient antibody transfer to the fetus. Further, the cord:maternal transfer ratios of RSV vaccination 2-3 and 3-4 weeks prior to delivery were significantly lower (p=0.008 and p=0.03, respectively) than the transfer ratios for anti-pertussis antibodies generated from routine Tdap vaccination, which typically occurs prior to 30 weeks’ gestation (**Figure 4A**). In contrast, RSV vaccination 4-5 and >5 weeks prior to delivery had similar transfer ratios to those generated from pertussis vaccination.

**Figure 4:**
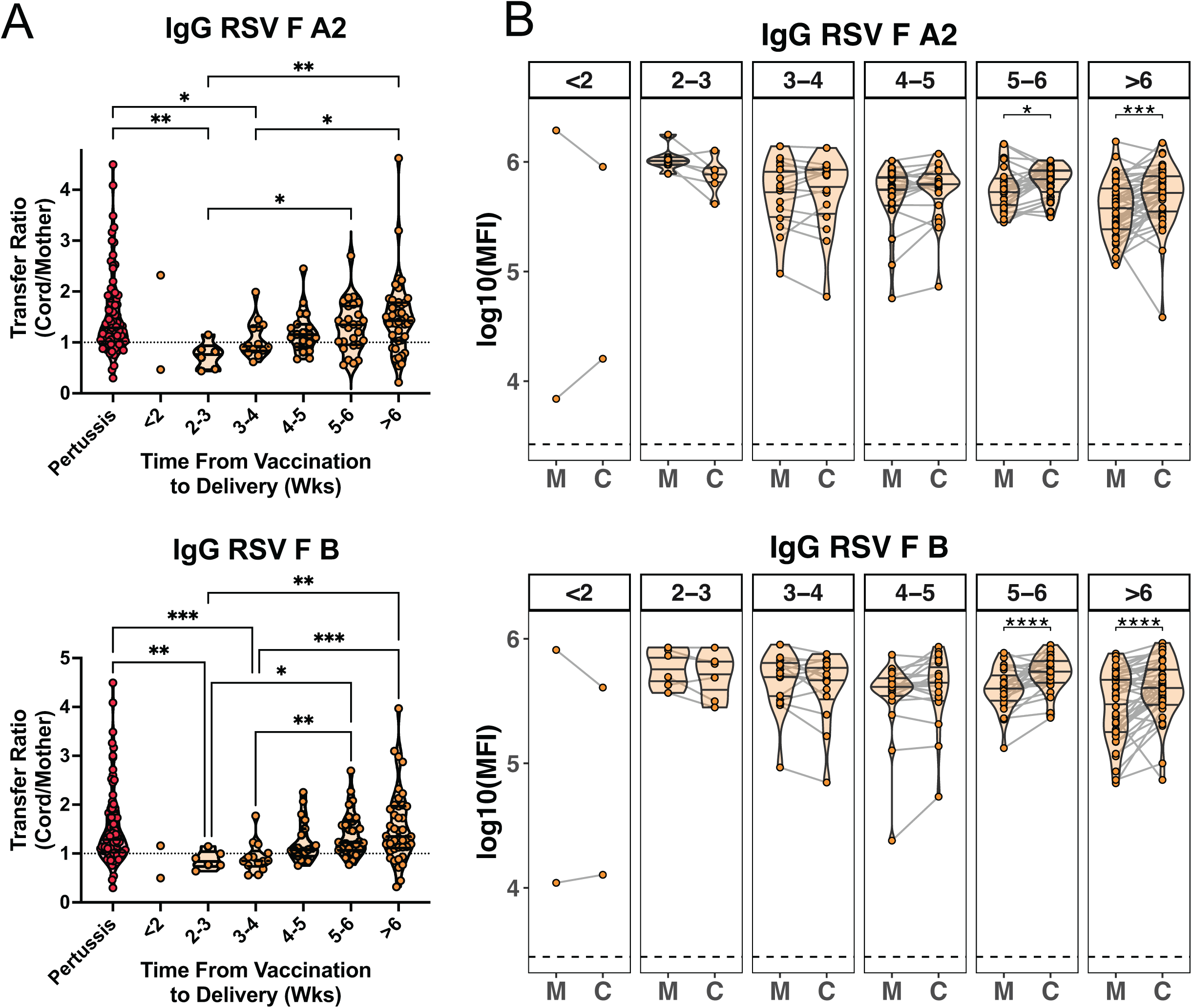
Transplacental transfer of maternal anti-F RSV IgG by weeks elapsed from RSV vaccination to delivery. The violin dot plots show the **(A)** transfer ratio (cord:maternal IgG) for antibodies against RSV Fusion (F) A2 (**top**) and RSV F B (**bottom**) stratified by weeks elapsed from vaccination to delivery. These transfer ratios were compared to those for anti-pertussis antibodies in dyads where the mother received the Tetanus, Diphtheria, Pertussis (Tdap) vaccine at < 30 weeks gestational age. The dashed line indicates a transfer ratio of 1 (definition of efficient transfer, or equivalent antibody levels in the umbilical cord and maternal compartments). Differences between the groups were assessed by Kruskal-Wallis test, followed by Dunn’s post hoc test. P values were further corrected using the Benjamini-Hochberg procedure (\**p*<0.05, \*\*\*\**p*<0.0001). **(B)** The violin dot plots represent levels of IgG against RSV F A2 (top) and RSV F B (bottom) in maternal (M) and cord (C) samples, stratified by weeks elapsed from RSV vaccination to delivery (weeks elapsed = numbers in box header). Lines represent maternal-cord dyads. Dashed line represents the median + 3 standard deviations above PBS background levels. Significance between groups was assessed by Wilcoxon-signed rank test (\**p*<0.05, \*\**p*<0.01, \*\*\**p*<0.001, \*\*\*\**p*<0.0001).

Plotting individual mother-cord dyads by weeks elapsed between vaccination and delivery further demonstrated that the expected significant increase in cord relative to maternal levels of anti-RSV F antibodies only occurred when the vaccine was administered ≥5 weeks prior to delivery (**Figure 4B**). Maternal, cord, and 2-month old infant anti-RSV F antibody levels by weeks elapsed from vaccination to delivery are depicted in **Supplementary** Figure 2A-C and demonstrate a significant impact of timing of RSV vaccination on maternal antibody levels, which are highest 2-3 weeks after vaccination, and require time to transit transplacentally to the umbilical cord, as reflected in the higher cord:maternal transfer ratios observed > 5 weeks post-vaccination.

We applied a linear mixed effects model to examine whether gestational age at vaccination was an independent factor in the efficient transplacental transfer of maternal RSV-specific IgG. For anti-RSV F A2, weeks elapsed from vaccination to delivery significantly associated with transplacental transfer efficiency, as measured by cord:maternal IgG ratios, while gestational age at vaccination did not (**Table 2**). For anti-RSV F against B strain, however, both weeks elapsed from vaccination to delivery and gestational age at vaccination were significantly associated with transplacental transfer efficiency of vaccine-elicited maternal IgG (**Table 3**). There was no statistically significant interaction between these two variables for either strain.

**Table 2:** Linear mixed-effects model of association between vaccine timing and placental transfer efficiency of anti-RSV-F A2 IgG. Weeks elapsed from vaccination to delivery has a significant impact in the placental transfer efficiency (measured by cord: maternal transfer ratio) of anti-RSV-F A2 IgG (p=0.0129). Gestational age at RSV vaccination did not have a significant impact (p=0.1299). GA of RSV = Gestational age at RSV vaccination (weeks); Weeks Elapsed from Vax to Delivery= number of weeks elapsed from vaccination to delivery. ^a^Likelihood Ratio Test (LRT) comparing Weeks Elapsed from Vaccination to Delivery model with Intercept Only model. ^b^LRT comparing GA of RSV model with Intercept Only model. ^c^LRT comparing Weeks Elapsed from Vaccination to Delivery + GA of RSV model with Weeks Elapsed from Vaccination to Delivery Only model. ^d^LRT comparing Weeks Elapsed from Vaccination to Delivery + GA of RSV + (Weeks Elapsed from Vaccination to Delivery * GA of RSV) model with Weeks Elapsed from Vaccination to Delivery + GA of RSV model.

| Model | -2 Log Likelihood | Likelihood Ratio Test (LRT) Statistic (G) | Degrees of Freedom | P-value |
| --- | --- | --- | --- | --- |
| Intercept Only | -97.15721 |  |  |  |
| Weeks Elapsed from Vax to Delivery<sup>a</sup> | -90.81157 | 12.69128<sup>a</sup> | 6 | 0.0129 (*) |
| GA of RSV<sup>b</sup> | -93.60024 | 7.113928<sup>b</sup> | 6 | 0.1299 (ns) |
| Weeks Elapsed from Vax to Delivery + GA of RSV<sup>c</sup> | -89.53744 | 2.548261<sup>c</sup> | 10 | 0.6360 (ns) |
| Weeks Elapsed from Vax to Delivery + GA of RSV + (Weeks Elapsed from Vax to Delivery) * (GA of RSV)<sup>d</sup> | -86.06822 | 6.938445<sup>d</sup> | 20 | 0.7312 (ns) |

**Table 3:**
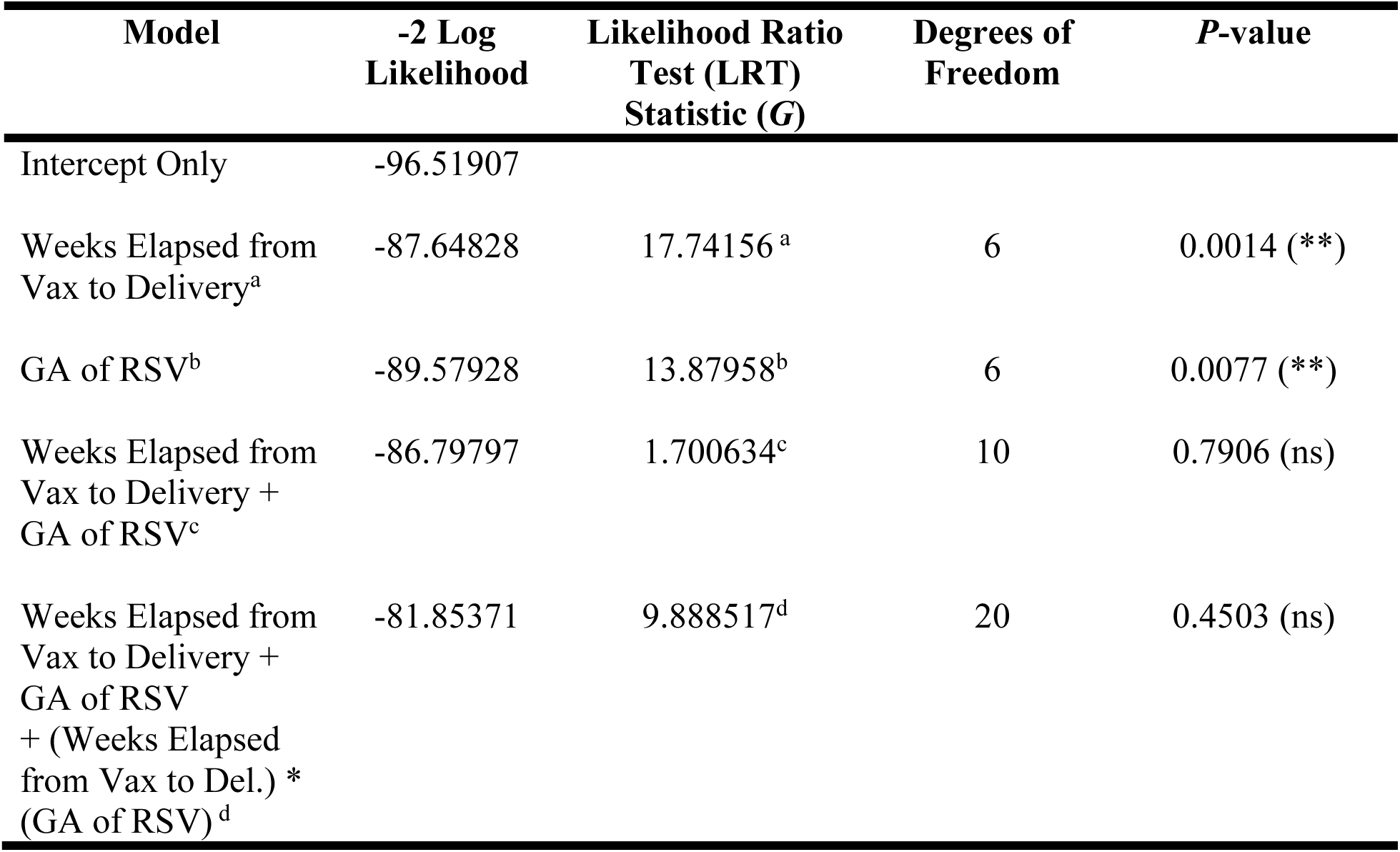
Linear mixed-effects model of association between vaccine timing and placental transfer efficiency of anti-RSV-F B IgG. Weeks elapsed from vaccination to delivery has a significant impact in the transfer ratio of anti-RSV-F B IgG of maternal-cord dyads (p=0.0014). Gestational age at RSV vaccination also has significant impact (p=0.0077). GA of RSV = Gestational age at RSV vaccination (weeks); Weeks Elapsed from Vax to Del.= number of weeks elapsed from vaccination to delivery. ^a^Likelihood Ratio Test (LRT) comparing Weeks Elapsed from Vaccination to Delivery model with Intercept Only model. ^b^LRT comparing GA of RSV model with Intercept Only model. ^c^LRT comparing Weeks Elapsed from Vaccination to Delivery + GA of RSV model with Weeks Elapsed from Vaccination to Delivery Only model. ^d^LRT comparing Weeks Elapsed from Vaccination to Delivery + GA of RSV + (Weeks Elapsed from Vaccination to Delivery * GA of RSV) model with Weeks Elapsed from Vaccination to Delivery + GA of RSV model.

| Model | -2 Log Likelihood | Likelihood Ratio Test (LRT) Statistic (G) | Degrees of Freedom | P-value |
| --- | --- | --- | --- | --- |
| Intercept Only | -96.51907 |  |  |  |
| Weeks Elapsed from Vax to Delivery<sup>a</sup> | -87.64828 | 17.74156<sup>a</sup> | 6 | 0.0014 (**) |
| GA of RSV<sup>b</sup> | -89.57928 | 13.87958<sup>b</sup> | 6 | 0.0077 (**) |
| Weeks Elapsed from Vax to Delivery + GA of RSV<sup>c</sup> | -86.79797 | 1.700634<sup>c</sup> | 10 | 0.7906 (ns) |
| Weeks Elapsed from Vax to Delivery + GA of RSV + (Weeks Elapsed from Vax to Del.) * (GA of RSV)<sup>d</sup> | -81.85371 | 9.888517<sup>d</sup> | 20 | 0.4503 (ns) |

## Comment

### Principal Findings

Here we demonstrate in a cohort of 142 individuals that maternal RSV vaccination with bivalent preFusion protein (Abrysvo, Pfizer) at least 5 weeks prior to delivery is associated with optimal transplacental transfer of maternal anti-RSV IgG. As RSV-specific antibody levels at birth have been demonstrated to be correlates of protection against RSV infection in infants too young to yet to receive their own vaccines,^24–26^ maternal RSV vaccination earlier in the approved 32-36 week window (to maximize the likelihood that at least 5 weeks elapse between vaccination and delivery) may confer the best possible protection for the neonate and infant. This finding has important implications for counseling pregnant patients about vaccine timing; currently vaccination any time within the 32-36 week approved window is presumed to be equivalent. This finding also informs future evaluations of vaccine timing, as a recent study demonstrated no association between nonadjuvanted bivalent RSV preF vaccination and preterm birth in a cohort study of 2973 patients who delivered during the 2023 to 2024 RSV vaccine season at two New York City hospitals.^13^

We also found that RSV vaccination from 32-36 weeks’ gestation generated significantly higher anti-F antibody levels than did natural RSV infection, and that the Abrysvo vaccine does not boost anti-RSV G antibody levels, raising the possibility of using anti-RSV G antibodies as an internal marker of prior natural infection, just as antibodies against SARS-CoV-2 Nucleocapsid (N) were used to distinguish natural infection from vaccine-induced antibodies. Finally, we demonstrated that anti-F antibodies derived from vaccination have significantly greater persistence in 2-month-old infants than do anti-G antibodies, which presumably reflect prior natural infection.

The greater durability of anti-F antibodies compared to naturally-elicited anti-RSV antibodies highlights the importance of maternal vaccination in neonatal and infant protection.

### Results in the Context of What is Known

Concerns about possible vaccine-associated preterm birth drove the FDA and ACIP’s approval of the Abrysvo RSV vaccine in pregnancy for administration only between 32+0 and 36+6 weeks’ gestation. This approval was based on limited data from 45% of the Phase 3 randomized controlled trial, and 38% of the Phase 2b randomized controlled trial, who received vaccine or placebo during the 32-36 week dosing interval.^6,27^ Prior work by our group examining infant protection from the COVID-19 mRNA vaccines found that vaccination later in the 32-36 week window was associated with lower 6-month infant IgG levels than vaccination earlier in the 32-36 week window.^18^ Our results, in line with this prior work,^18^ point to the need to examine transplacental antibody transfer week-by-week within this approved vaccine window.

Our finding that maternal RSV vaccination at least 5 weeks prior to delivery was associated with optimal transplacental transfer of maternal anti-RSV IgG is consistent with studies in Tdap vaccinees demonstrating higher antibody levels in the cord blood of neonates after maternal immunization at 27-30+6 weeks, compared to maternal immunization at >31 weeks’ gestation.^12,28,29^ These findings suggest that efficient transplacental transfer of maternal antibody following any vaccination requires at least 30 days from vaccination to delivery.^30^ We identified that transplacental transfer efficiency for anti-RSV F IgG was non-inferior (not significantly lower) than that resulting from pertussis vaccination prior to 30 weeks even at 4 weeks post-vaccination, although vaccination ≥ 5 weeks prior to delivery was associated with the best transplacental transfer.

Our group has previously demonstrated in a geographically-diverse 300 person cohort that antibodies to RSV-F and -G antigens from naturally-occurring infection were nearly ubiquitous in pregnant individuals and their neonates,^19^ but the significantly enhanced infant protection conferred by maternal vaccination^6,9^ suggests key differences in antibody titer and/or quality generated by vaccination. Here we found that vaccine-induced antibody titers are higher than those induced by natural infection and persist longer in infants. These findings are consistent with our findings for anti-Spike antibodies induced by COVID-19 vaccination versus natural infection in mother-infant dyads and 6-month infants.^16,17^ Multi-level linear regression models demonstrated a gestational-age-specific impact of vaccination on transplacental transfer for B strain only, suggesting additional evaluation is needed in larger cohorts to determine if gestational age within the 32-36 week window exerts a further effect on transplacental antibody transfer.

### Clinical Implications

As cord blood and infant antibody levels after maternal vaccination in pregnancy have been demonstrated to be important correlates of infant protection from respiratory illnesses,^16,18,31,32^ our results provide a strong rationale for changing how we counsel pregnant patients about receiving the RSV vaccine within the approved window, and reconsidering the current timing recommendations as real-world vaccine safety and efficacy data continue to emerge.^13^ Given that preterm infants account for 25% of RSV related hospitalizations,^5^ vaccinating earlier in the 32-36 week window or expanding the window may have greatest benefit for infants born to mothers with spontaneous preterm birth, often an unpredictable event. Our work raises the question of whether indications for the monoclonal antibody nirsevimab postnatally should be expanded to infants born beyond the 2-week post-vaccination window, which would require much larger studies to determine more precisely what antibody levels are required for optimal protection.

### Research Implications

This work highlights the power of detailed antibody analyses to provide clinically useful information. Given insights into transplacental transfer and immune cell activation in the infant provided by analyses of canonical (FcRn) and non-canonical (FcγR1-3) Fc-receptor binding properties of antibodies,^14,15,18,22^ future work should evaluate FcR binding, antibody functions, and the glycan profile of vaccine-generated antibodies in maternal, cord and infant blood through 6 months of age. Understanding the potential additive protection for infants provided by breastmilk from RSV-vaccinated mothers and characterization of the contribution of maternal vaccination to infant mucosal immunity against RSV is an important area for future research. Finally, evaluating the impact of neonatal and infant sex on antibody properties and transplacental transfer is an important future direction, given the known impact of fetal sex on maternal and infant immunity.^33–35^ This work also has the potential to provide insights into maternal vaccination relevant far beyond RSV vaccination itself, informing development of rational vaccines for maternal and infant protection and definitions of optimal vaccination strategy.

### Strengths and Limitations

A strength of this study is the relatively large and diverse cohort enrolled across two academic medical centers in New York and Boston. Timing of Tdap vaccination was also known precisely, facilitating biological comparisons between transfer of antibody from a vaccine administered 27-32 weeks versus 32-36 weeks. This group also had no possible prior exposure to RSV vaccine, which will not be the case for future cohorts. A further strength is the use of two biological comparator groups (1) maternal-infant dyads with prior natural RSV infection, and (2) maternal-infant dyads with Tdap-generated antibodies.

Despite inclusion of a relatively large cohort, one limitation is the relatively small numbers of participants for week-over-week analysis, precluding analysis of the impact of fetal and infant sex. A larger future cohort might also permit discrimination between the relative effects of gestational age at vaccination versus time elapsed from vaccination to delivery. Breastmilk antibodies also remain a critical and understudied area with respect to RSV vaccination, especially the potentially additive or synergistic protection these antibodies could confer. Finally, while antibodies are correlates of protection, determining whether the more efficient transfer of antibodies will lead to enhanced protection for infants through 2 and 6 months of age will require larger studies.

### Conclusions

Our results suggest that maternal RSV vaccination at least 5 weeks prior to delivery is associated with optimal RSV-specific IgG transplacental transfer. Additional larger studies are needed to further refine these findings and their clinical implications, particularly whether the timing indication for monoclonal antibody should be expanded beyond infants who delivered within 2 weeks of maternal RSV vaccination.

## Data Availability

All data produced in the present study are available upon reasonable request to the authors

## References

1 Prevention, C. f. D. C. a. Respiratory Syncytial Virus Infection-RSV Surveillance and Research, 2024).

2 Li, Y. et al. Global, regional, and national disease burden estimates of acute lower respiratory infections due to respiratory syncytial virus in children younger than 5 years in 2019: a systematic analysis. Lancet 399, 2047–2064, doi:10.1016/S0140-6736(22)00478-0 (2022).

3 McLaughlin, J. M. et al. Respiratory Syncytial Virus-Associated Hospitalization Rates among US Infants: A Systematic Review and Meta-Analysis. J Infect Dis 225, 1100–1111, doi:10.1093/infdis/jiaa752 (2022).

4 Fleming-Dutra, K. E. et al. Use of the Pfizer Respiratory Syncytial Virus Vaccine During Pregnancy for the Prevention of Respiratory Syncytial Virus-Associated Lower Respiratory Tract Disease in Infants: Recommendations of the Advisory Committee on Immunization Practices - United States, 2023. MMWR Morb Mortal Wkly Rep 72, 1115–1122, doi:10.15585/mmwr.mm7241e1 (2023).

5 Wang, X. et al. Global disease burden of and risk factors for acute lower respiratory infections caused by respiratory syncytial virus in preterm infants and young children in 2019: a systematic review and meta-analysis of aggregated and individual participant data. Lancet 403, 1241–1253, doi:10.1016/S0140-6736(24)00138-7 (2024).

6 Kampmann, B. et al. Bivalent Prefusion F Vaccine in Pregnancy to Prevent RSV Illness in Infants. New England Journal of Medicine 388, 1451–1464, doi:10.1056/NEJMoa2216480 (2023).

7 CDC recommends new vaccine to help protect babies against severe respiratory syncytial virus (RSV) illness after birth, https://www.cdc.gov/media/releases/2023/p0922-RSV-maternal-vaccine.html> (2023).

8 Administration, F. a. D. in Center for Biologics Evaluation and Research, VRBPAC meeting.

9 Dieussaert, I. et al. RSV Prefusion F Protein-Based Maternal Vaccine - Preterm Birth and Other Outcomes. N Engl J Med 390, 1009–1021, doi:10.1056/NEJMoa2305478 (2024).

10 Jones, J. M. et al. Use of Nirsevimab for the Prevention of Respiratory Syncytial Virus Disease Among Infants and Young Children: Recommendations of the Advisory Committee on Immunization Practices - United States, 2023. MMWR Morb Mortal Wkly Rep 72, 920–925, doi:10.15585/mmwr.mm7234a4 (2023).

11 ACOG Practice Advisory; Ault K.A. H. B. L., Riley, L.E. Maternal Respiratory Syncytial Virus Vaccination, <https://www.acog.org/clinical/clinical-guidance/practice-advisory/articles/2023/09/maternal-respiratory-syncytial-virus-vaccination#:~:text=On%20August%203%2C%202023%2C%20the,entering—their%20first%20RSV%20season.> (2023).

12 Society for Maternal-Fetal Medicine. Electronic address, p. s. o., Joseph, N. T., Kuller, J. A., Louis, J. M. & Hughes, B. L. Society for Maternal-Fetal Medicine Statement: Clinical considerations for the prevention of respiratory syncytial virus disease in infants. Am J Obstet Gynecol 230, B41–B49, doi:10.1016/j.ajog.2023.10.046 (2024).

13 Son, M. et al. Nonadjuvanted Bivalent Respiratory Syncytial Virus Vaccination and Perinatal Outcomes. JAMA Netw Open 7, e2419268, doi:10.1001/jamanetworkopen.2024.19268 (2024).

14 Atyeo, C. et al. COVID-19 mRNA vaccines drive differential antibody Fc-functional profiles in pregnant, lactating, and nonpregnant women. Sci Transl Med 13, eabi8631, doi:10.1126/scitranslmed.abi8631 (2021).

15 Atyeo, C. G. et al. Maternal immune response and placental antibody transfer after COVID-19 vaccination across trimester and platforms. Nat Commun 13, 3571, doi:10.1038/s41467-022-31169-8 (2022).

16 Shook, L. L. et al. Durability of Anti-Spike Antibodies in Infants After Maternal COVID-19 Vaccination or Natural Infection. JAMA 327, 1087–1089, doi:10.1001/jama.2022.1206 (2022).

17 Gray, K. J. et al. Coronavirus disease 2019 vaccine response in pregnant and lactating women: a cohort study. Am J Obstet Gynecol 225, 303 e301–303 e317, doi:10.1016/j.ajog.2021.03.023 (2021).

18 Lopez, P. A. et al. Placental transfer dynamics and durability of maternal COVID-19 vaccine-induced antibodies in infants. iScience 27, 109273, doi:10.1016/j.isci.2024.109273 (2024).

19 Edlow, A. G. L. Z., P.A.; Shook, L.L.; Jasset, O.; Gray, K.J.; Lauffenburger, D.L.; Elovitz, M.A. in *Society for Maternal-Fetal Medicine 2024 Annual Meeting* (National Harbor, Maryland, February 14, 2024, 2023).

20 Tomaras, G. D. et al. Initial B-cell responses to transmitted human immunodeficiency virus type 1: virion-binding immunoglobulin M (IgM) and IgG antibodies followed by plasma anti-gp41 antibodies with ineffective control of initial viremia. J Virol 82, 12449–12463, doi:10.1128/JVI.01708-08 (2008).

21 Martinez, D. R. et al. Fc Characteristics Mediate Selective Placental Transfer of IgG in HIV-Infected Women. Cell 178, 190–201 e111, doi:10.1016/j.cell.2019.05.046 (2019).

22 Atyeo, C. et al. Compromised SARS-CoV-2-specific placental antibody transfer. Cell 184, 628–642 e610, doi:10.1016/j.cell.2020.12.027 (2021).

23 Clements, T. et al. Update on Transplacental Transfer of IgG Subclasses: Impact of Maternal and Fetal Factors. Front Immunol 11, 1920, doi:10.3389/fimmu.2020.01920 (2020).

24 Buchwald, A. G. et al. Respiratory Syncytial Virus (RSV) Neutralizing Antibodies at Birth Predict Protection from RSV Illness in Infants in the First 3 Months of Life. Clin Infect Dis 73, e4421–e4427, doi:10.1093/cid/ciaa648 (2021).

25 Fong, Y. et al. Antibody Correlates of Protection From Severe Respiratory Syncytial Virus Disease in a Vaccine Efficacy Trial. Open Forum Infect Dis 10, ofac693, doi:10.1093/ofid/ofac693 (2023).

26 Glezen, W. P., Paredes, A., Allison, J. E., Taber, L. H. & Frank, A. L. Risk of respiratory syncytial virus infection for infants from low-income families in relationship to age, sex, ethnic group, and maternal antibody level. J Pediatr 98, 708–715, doi:10.1016/s0022-3476(81)80829-3 (1981).

27 Fleming-Dutra, K. E. Evidence to Recommendations Framework Updates Pfizer Maternal RSVpreF Vaccine. Centers for Disease Control and Prevention, ACIP General Meeting *September 22*, 2023 https://www.cdc.gov/vaccines/acip/meetings/downloads/slides-2023-09-22/06-Mat-Peds-Fleming-Dutra-508.pdf (2023).

28 Abu Raya, B., et al. The effect of timing of maternal tetanus, diphtheria, and acellular pertussis (Tdap) immunization during pregnancy on newborn pertussis antibody levels - a prospective study. Vaccine 32, 5787–5793, doi:10.1016/j.vaccine.2014.08.038 (2014).

29 Abu Raya, B., et al. Immunization of pregnant women against pertussis: the effect of timing on antibody avidity. Vaccine 33, 1948–1952, doi:10.1016/j.vaccine.2015.02.059 (2015).

30 van den Berg, J. P., Westerbeek, E. A., van der Klis, F. R., Berbers, G. A. & van Elburg, R. M. Transplacental transport of IgG antibodies to preterm infants: a review of the literature. Early Hum Dev 87, 67–72, doi:10.1016/j.earlhumdev.2010.11.003 (2011).

31 Halasa, N. B. et al. Effectiveness of Maternal Vaccination with mRNA COVID-19 Vaccine During Pregnancy Against COVID-19-Associated Hospitalization in Infants Aged <6 Months - 17 States, July 2021-January 2022. MMWR Morb Mortal Wkly Rep 71, 264–270, doi:10.15585/mmwr.mm7107e3 (2022).

32 Halasa, N. B. et al. Maternal Vaccination and Risk of Hospitalization for Covid-19 among Infants. N Engl J Med 387, 109–119, doi:10.1056/NEJMoa2204399 (2022).

33 Bordt, E. A. et al. Maternal SARS-CoV-2 infection elicits sexually dimorphic placental immune responses. Sci Transl Med 13, eabi7428, doi:10.1126/scitranslmed.abi7428 (2021).

34 Klein, S. L. & Flanagan, K. L. Sex differences in immune responses. Nat Rev Immunol 16, 626–638, doi:10.1038/nri.2016.90 (2016).

35 Baines, K. J. & West, R. C. Sex differences in innate and adaptive immunity impact fetal, placental, and maternal healthdagger. Biol Reprod 109, 256–270, doi:10.1093/biolre/ioad072 (2023).

