## Supplementary Material for "Longer interval between maternal RSV vaccination and birth increases placental transfer efficiency"

### 1. Supplementary Methods

***Antibody quantification: Binding Antibody Multiplex Assay (BAMA)***

RSV and Pertussis toxin antigens were conjugated to MagPlex microspheres (Luminex) as previously described[20,21]. The antigens conjugated to the MagPlex microspheres are as follows: Pertussis Toxin (Sigma-Aldrich, Cat# P7208), RSV-F Strain A2, RSV-G Strain A2, RSV-F Strain 18537(B), RSV-G Strain 18537(B). Conjugated microspheres were incubated on 384-well black Aurora plate (Millipore Sigma, Cat# ABB2-00160A) for approximately 30 minutes before standards, controls and plasma samples were added. The standards, which included Pertussis Antiserum (NIBSC, WHO 06/140), Synagis/Palivizumab (Arexis AB, Sobi, Waltham, MA) and Human Reference Immune Globulin to RSV (BEI, Cat#NR-21973), were diluted in assay diluent (1% dry milk + 5% goat serum + 0.05% tween-20 in 1X phosphate buffer saline, pH 7.4.) at starting concentrations of 10IU/mL, 10ug/mL, 250ug/mL respectively and serially diluted 2- or 3- fold (Pertussis Antiserum, Synagis, Human Reference Immune Globulin to RSV, respectively). Additional controls, which included: (1) Human Antiserum to RSV, High Control (BEI Resources, Cat#NR-4021), (2) Human Antiserum to RSV, Medium Control (BEI Resources, Cat#NR-4022), (3) Human Antiserum to RSV, Low Control (BEI Resources, Cat#NR-4023), (4) Human Reference Antiserum to RSV (BEI Resources, Cat#NR-4020) and (5) Human IgG-Depleted Serum (BEI Resources, Cat#NR-49447), were added undiluted. The plasma samples were diluted in assay diluent at a 1:400-point dilution. Beads and diluted samples were incubated for 30 minutes, then IgG binding was detected using a PE-conjugated mouse anti-human IgG

(Southern Biotech, Cat#9040-09) at 2 µg/mL. The beads were washed and acquired on an xMap Intelliflex instrument (Luminex), and IgG binding was expressed as median fluorescence intensity (MFI). To assess assay background, the MFI of binding to wells that did not contain beads or sample (blank wells) and non-specific binding of the samples to unconjugated blank beads were evaluated during assay analysis. An antigen-specific antibody response was considered positive if above the lower limit of detection, defined as the mean + 3SD of PBS blank well MFIs for each antigen. To check for consistency between assays, the EC50 and maximum MFI values of the standards was tracked by Levy-Jennings charts.

**2. Supplementary Figures 1 and 2**

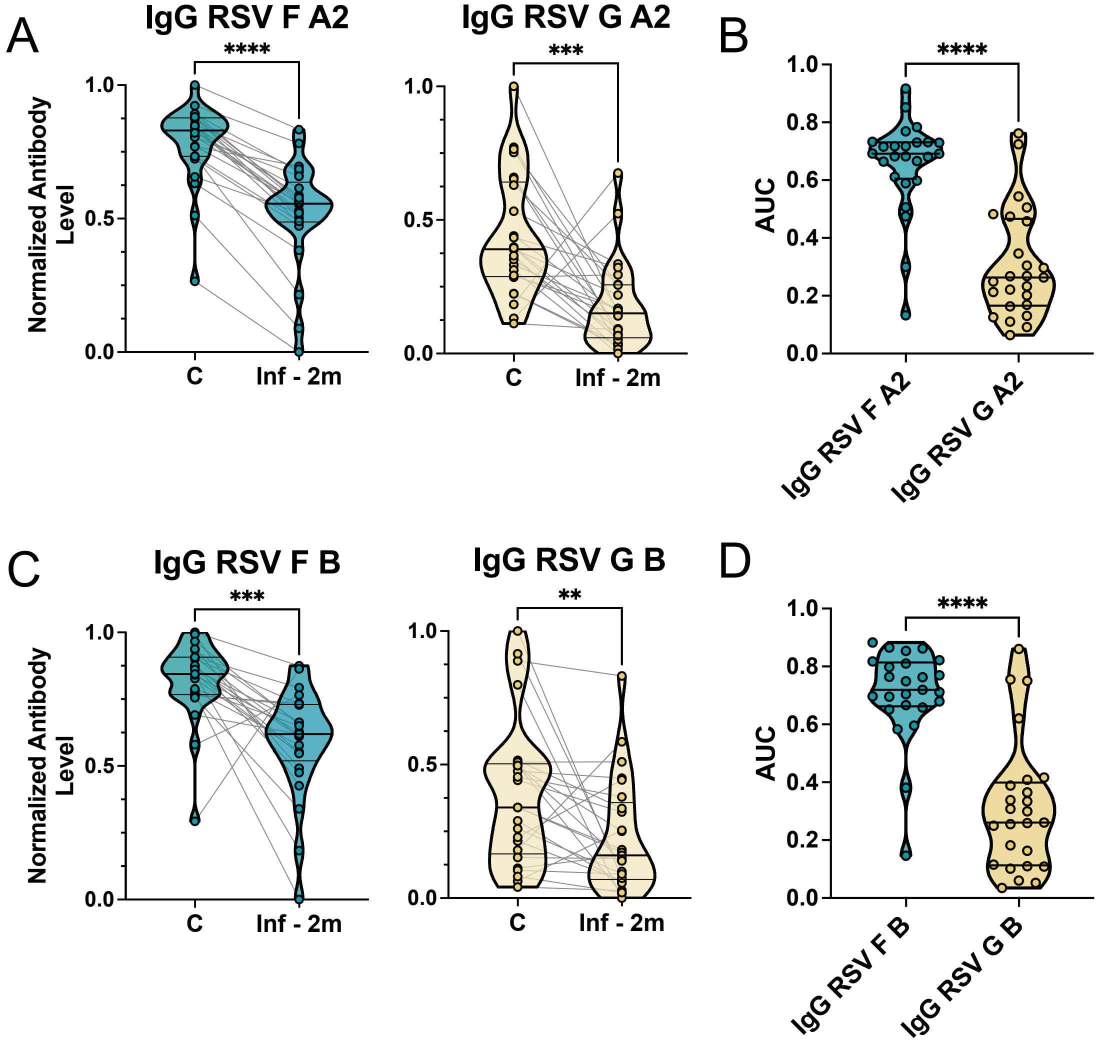


**Supplementary Figure 1**: **Evaluating persistence of anti-F and anti-G RSV IgG from birth to 2 months of age. (A)** Violin plots represent normalized IgG antibody levels targeted against RSV Fusion (F) A2 (Left) and G A2 (Right) in N=29 cord (C) and 2-month infants (Inf -2m), with lines connecting birth and postpartum samples from the same child. Significance between groups was assessed by Wilcoxon-signed rank test (****p*<0.001 ,*****p*<0.0001). **(B)** Violin plots represent IgG antibody persistence for anti-F A2 antibodies (from vaccination) versus anti-G A2 antibodies (from natural infection), defined by Area under the Curve (AUC) from birth to 2 months. AUC was calculated by taking the area beneath the matched cord-infant dyad line. Differences in AUC were assessed by Wilcoxon rank-sum test (*****p*<0.0001). **(C)** Violin plots represent normalized IgG antibody levels targeted against RSV F B (Left) and RSV G B (Right) in N=29 cord (C) and 2-month infants (Inf -2m), with lines connecting birth and postpartum samples from the same child. Significance between groups was assessed by Wilcoxon-signed rank test (****p*<0.001 ,***p*<0.01). **(D)** Violin plots represent IgG antibody persistence for anti-F B antibodies (from vaccination) versus anti-G B antibodies (from natural infection), defined by Area under the Curve (AUC) from birth to 2 months. Differences in AUC were assessed by Wilcoxon rank-sum test (*****p*<0.0001).

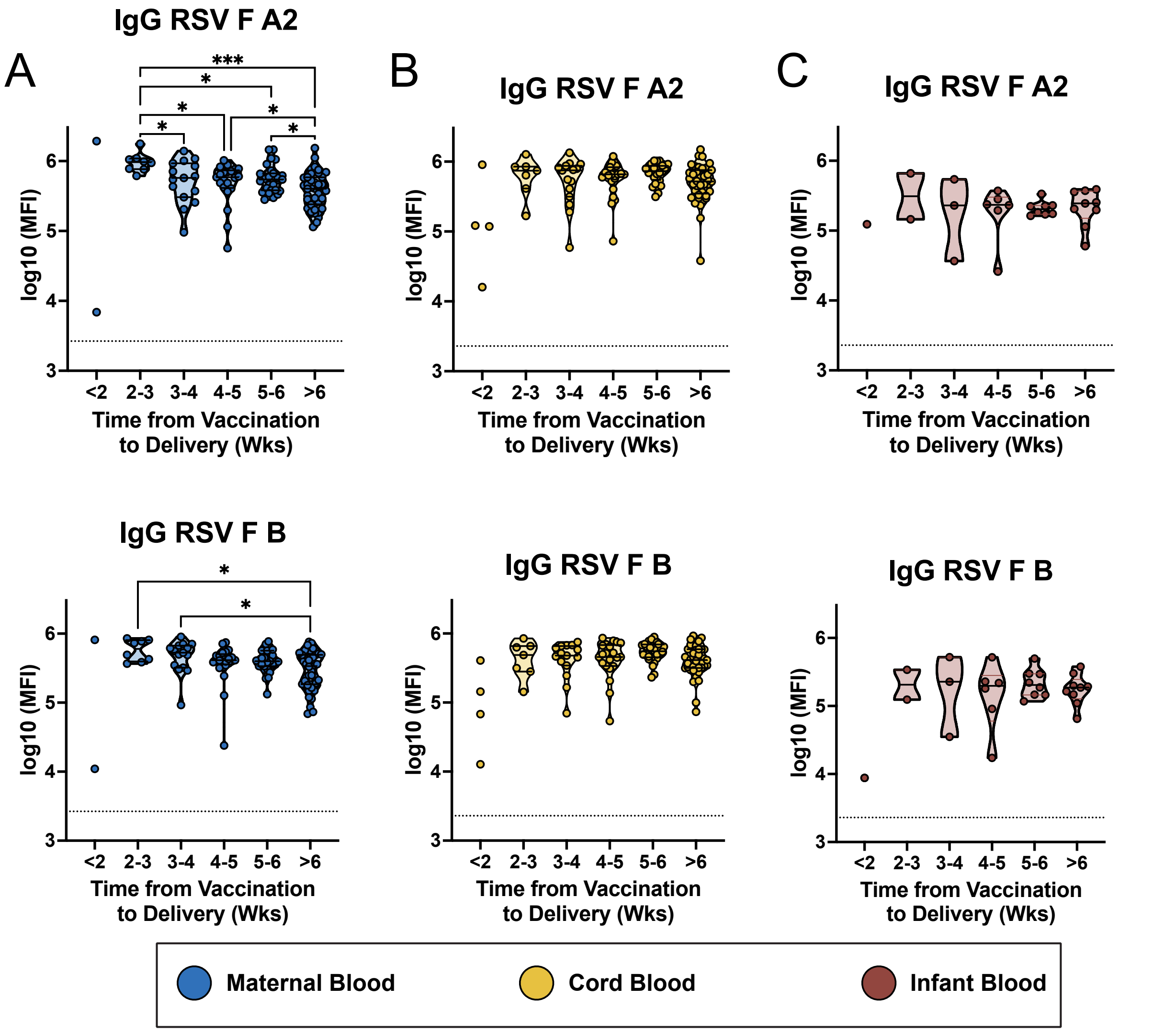


**Supplementary Figure 2**: **Maternal, umbilical cord, and 2-month infant anti-RSV F IgG levels by weeks elapsed from maternal vaccination**. Violin plots show the levels of IgG against RSV Fusion A2 (**top**) and B (**bottom**), stratified by the number of weeks elapsed from vaccination in **(A)** maternal, **(B)** cord blood, and **(C)** 2-month old infant samples. The dashed line indicates the threshold of detectability, defined as the median + 3 standard deviations above PBS background level. Differences between the groups were assessed by Kruskal-Wallis test, followed by Dunn's post hoc analysis if relevant. P values were further corrected for multiple comparisons using Benjamini-Hochberg procedure (*$p<0.05$, ***$p<0.001$).
